# Time delay epidemic model for COVID-19

**DOI:** 10.1101/2020.05.23.20111500

**Authors:** Montri Maleewong

**Affiliations:** Department of Mathematics, Faculty of Science, Kasetsart University, Bangkok, 10900, Thailand

**Keywords:** COVID-19, SIR, time delay, Monte Carlo, reproduction number

## Abstract

A time delay epidemic model is presented for the spread of the Coronavirus 2019 (COVID-19) in China. The time delay effects affect infected individuals. Monte Carlo simulation is performed to estimate the transmission and recovery rates. The basic reproduction number is estimated in terms of the average infected ratio. This ratio can be used to monitor the policy performance of disease control during the spread of the disease.

## 1 Introduction

The Coronavirus (COVID-19) is a new respiratory virus that has not been previously identified. It was reported first in Wuhan, Hubei province, China in December 2019. This virus probably originated from an animal but at present, it can spread from human to human. At this moment, it is unclear how easily this virus can spread among people. Understanding the spreading mechanism is a very important issue in the control of its extensive global spread. Mathematical modelling is a necessary tool to understand the transmission mechanism. Various research studies have been presented to describe the virus, see Anderson and May (1979), Diekmann and Heesterbeek (2000) and Xu and Ma (2009). To control the spread of the virus, a dimensionless constant called the basic reproduction number *R\** is usually estimated to predict the number of infected from susceptible individuals in the future. Basically *R\** ≈ 3 − 4 for influenza, but it is much larger for measles so that *R\** ≈ 16 − 18, see Keeling and Rohani (2008) and Anderson and May (1998). However, *R\** for COVID-19 is unclear, so in this work, we will apply the standard SIR model, Kermack and McKendrick (1927) to estimate *R\** and we will present an extension of the SIR model with two-time delays for infected individuals to obtain the best fit to the actual data using Monte Carlo Simulation. The transmission and recovery rates are approximated and a time delay effect is proposed. Finally, the infected ratio is defined and presented as an aid to monitoring control of the spread of the virus.

## 2 Epidemic models

The SIR model is a system of differential equations describing the spread of a disease in a population and is known as an epidemic model, see Kermack and McKendrick (1927). There are three “compartments” in the population of size *N*. Each compartment is a function of time *t* composed of the following: *S*(*t*) is the number of susceptible individuals not yet infected with the disease, *I*(*t*) is the number of infectious individuals, and *R*(*t*) is the number of recovery individuals from the disease who then have immunity.

Initially, the following assumptions are made to build the mathematical model.

- The populations of susceptible individuals and contagious infectives are very large so that random differences between individuals can be neglected.
- There are no natural births and deaths in this model. The disease is spread by contact from individual to individual.
- Individuals who recover from the disease have immunity.
- The population system is closed with no migration.

The system of differential equations describing the population change of each compartment (*S, I, R*) over time is expressed by

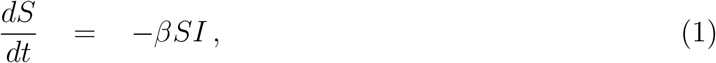

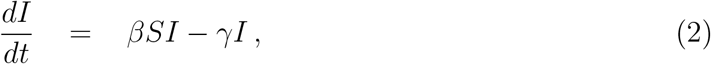

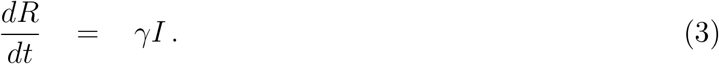

The SIR model describes the change of each compartment over time where *β* is the mean infection rate and *γ* is the mean recovery rate. The total number of individuals is *N = S* + *I* + *R*, and we make an important assumption that *N* is maintained over time. Note that model equations 1-3 are in dimensionless form where the number of each individual is divided by *N*.

The epidemic model described by the differential equations 1 - 3 has much studied from various viewpoints, see Kermack and McKendrick (1927). The main assumptions are that the mean infection and recovery rates are positive constants over time. In a real situation, these assumptions are not satisfied and they can be functions of time, or put in another way, they can be set as mean constants but the change for each compartment can be caused by a time-delay effect. For the present case of COVID-19, we propose a more general epidemic model which includes the time delay effect for both infected and recovered individuals. The implication is that the susceptible people are not immediately infected at any time when they are in contact with infected people but there should be a time-delay period of disease incubation. The time-delay effect can be applied to the recovery people meaning that the infected people do not immediately become recovery people at any time. Then the SIR model can be generalized with time delay effects as presented by the following.

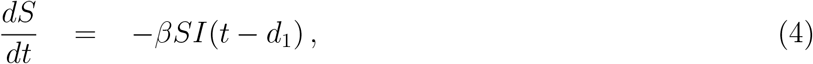

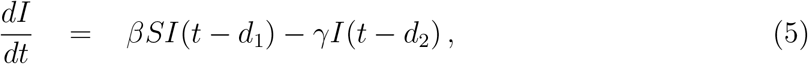

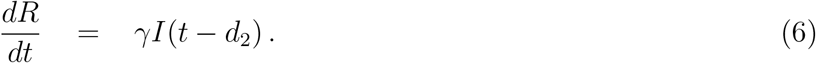

where *d*_1_ and *d_2_* represent the effects of time delay for the infective and recovered individuals, respectively.

## 3 Simulation results

We show the numerical results and compare them with real data collected by the Johns Hopkins Github repository and summarized again by Nadu (2020). A case study is considered in China since a complete data set is available for describing and fitting parameters in the present model. Three are three-time stages for infective individuals: the early stage where the number of cases is slowly increasing, the middle stage of a fast increase, and the final stage of decreasing to the steady-state. Data are used from January 22, 2020, to March 15, 2020. We assume the total number of people as *N* = 83,000. At the beginning of the study period *t*_0_, the initial numbers of susceptible, infective and recovery individuals are given by *S*_0_ = 82408/*N*, *I*_0_ = 547/*N*, and *R*_0_ = 45/*N*, respectively, according to the daily report on January 22, 2020. Before performing the simulation, plots of actual data for susceptible, infective and recovery individuals are shown in Figures 1 and 2. The number of people in the model is multiplied by *N* to compare with the actual data.

**Figure 1:**
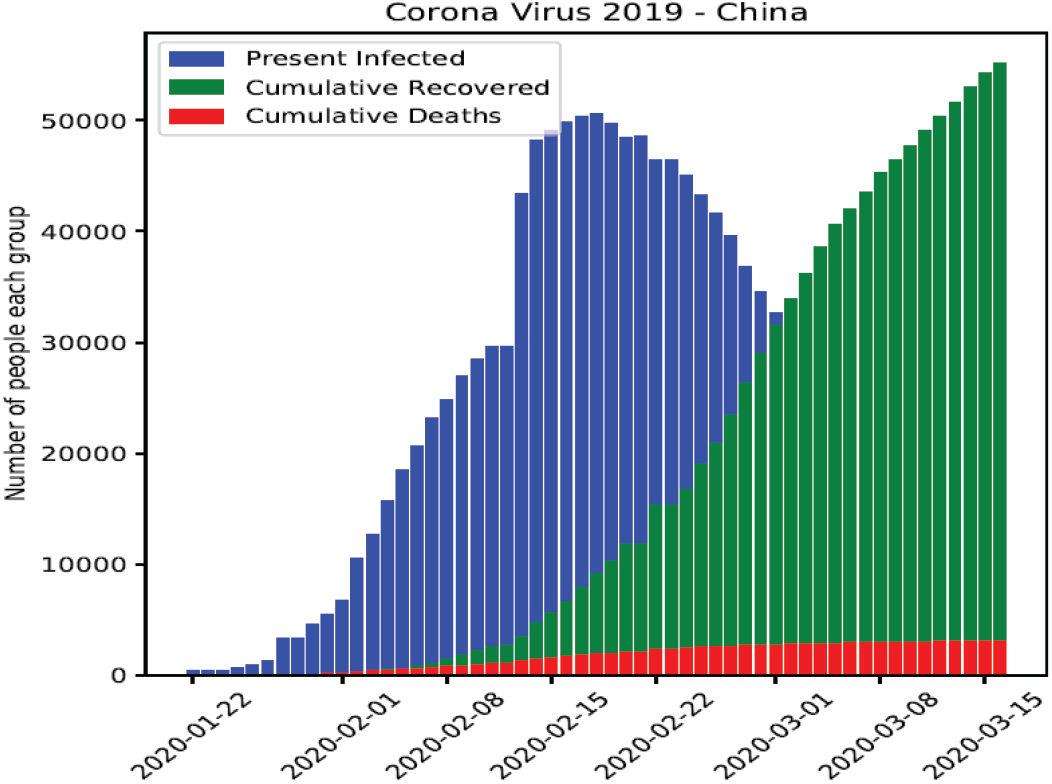
Bar plot of actual data for infected, recovery and death people due to COVID-19.

**Figure 2:**
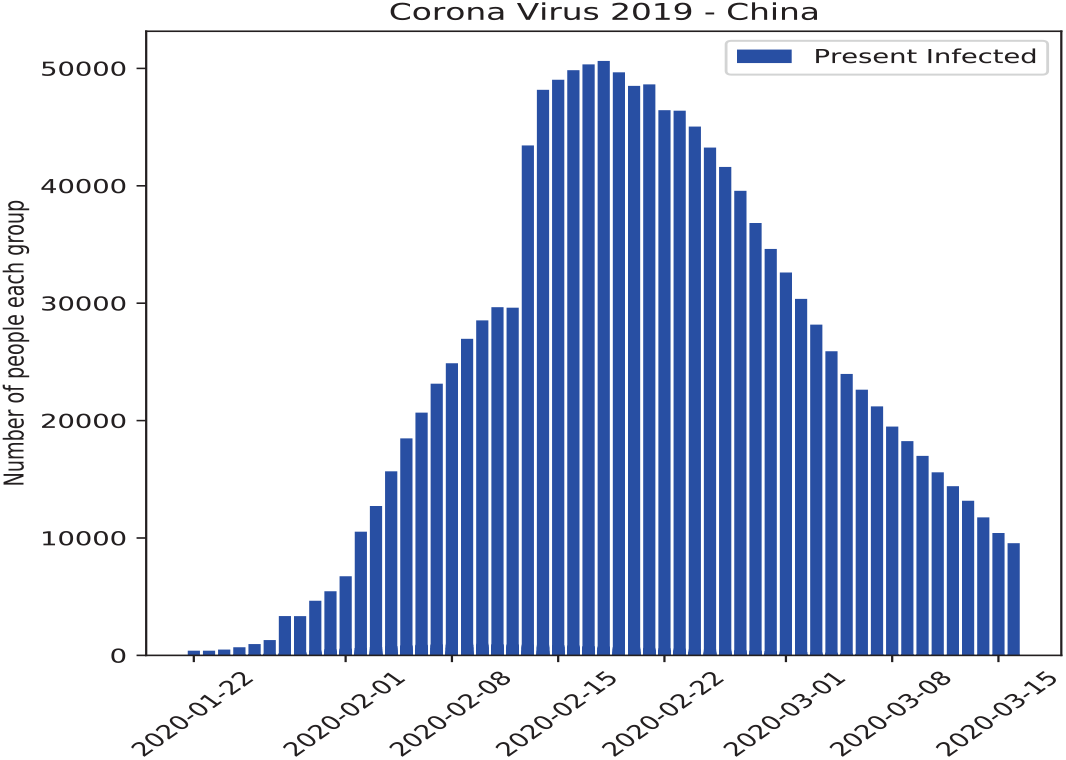
Bar plot of actual data for infected people due to COVID-19.

To numerically approximate the parameters *β* and *γ* in the standard SIR model in (1)-(3), we apply Monte Carlo simulation using 1,000 random iterative steps. The system ODEs can be integrated using the *ddeint* package, Zulko (2019). Two sets of uniformly distributed random number are generated to assign the guess values of *β* and *γ* for each iteration where we consider 0.1 *< β <* 0.6 and 0.02 *<* γ *<* 0.08. The optimal values are obtained by minimizing the percentage of the root mean square error (RMSE) between the actual data and the simulation result for infected individuals over 61 days. We obtain the minimum value of the percentage RMSE 6.438% corresponding to the optimal values *β* = 0.345 and 0.045. The comparison between the actual data and the standard SIR simulation is shown in Figure 3. If the basic reproduction number *R\** is defined by *βS*_0_*/γ*, we obtain *R\** = 7.61 which is relatively larger than the influenza spread *R\** = 3 − 4, see Keeling and Rohani (2008) and Anderson and May (1998). This case *R\** = 7.61 represents the COVID-19 in China that might be over-estimation since we use *S*_0_ to approximate *R^*^* and that might be accurate only in the early stage. Instead of using *S*_0_, we define 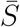 as the average of susceptible individuals over the whole model period, so we obtain S from the simulation as 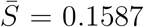, and thus 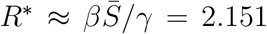. Again, this value might be an under-estimate since the simulation result in Figure 3 does not fit well with the actual data.

**Figure 3:**
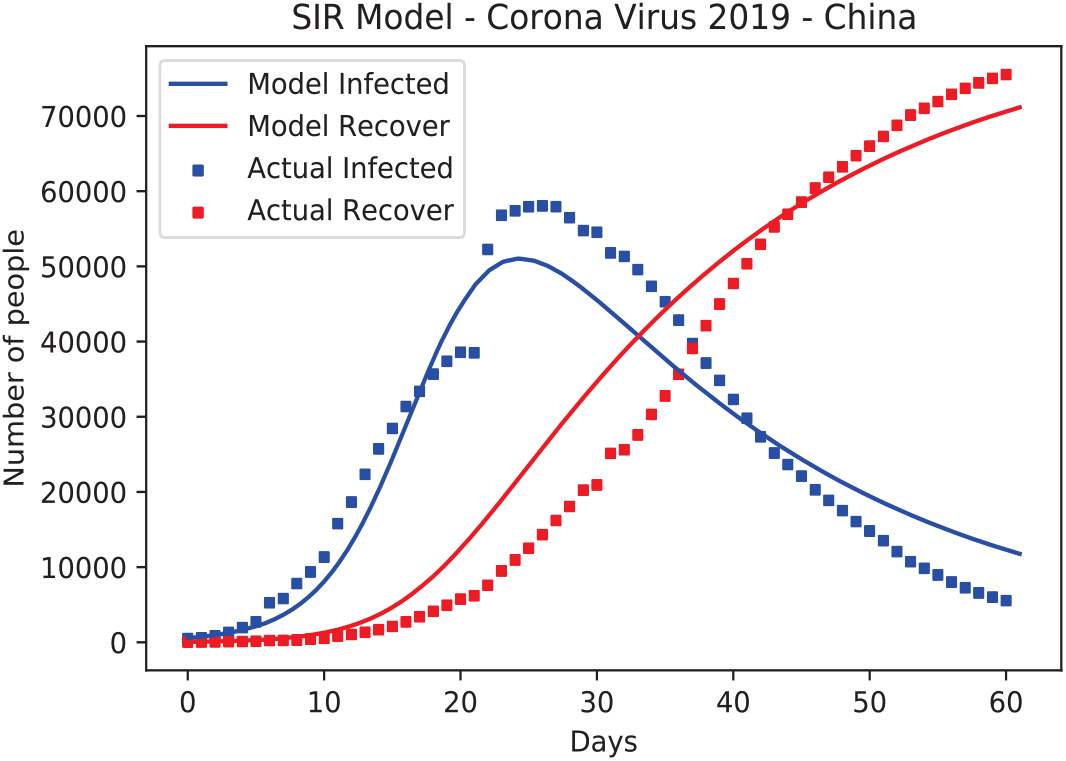
The results of SIR model

To obtain a better fit than the standard SIR model, the concept of two-time delay is proposed using equations (4)-(6) where *d*_1_ is the time delay effect from susceptible to infected individuals and *d*_2_ is the time delay effect from the infected to recovery individuals. To obtain the best fit, we apply the Monte Carlo simulation but here *d*_1_ and *d*_2_ are two additional parameters which must be considered in the fit as well. So, four set of uniformly distributed random numbers are generated to find the optimal values of *β*, *γ*, *d*_1_, and *d*_2_. Here, we specify the parameter ranges as: 1.5 ≤ *β* ≤ 2.3, 0.03 ≤ *γ* ≤ 0.08, 7 *< d*_1_ ≤ 14, and 0 *< d*_1_ ≤ 7. After performing a simulation using 1,000 iteration steps, we obtain the optimal values *β* = 1.747, *γ* = 0.05, *d*_1_ ≈ 12, *d*_2_ *≈* 5 with the percentage RMSE of 4.466%. The comparison between the actual data and the SIR with the time delay model is shown in Figure 4. It can be seen that the simulation results for both infected and recovery individuals agree well with the actual data. If we calculate the ratio, *βS*_0_*/γ* ≈ 34.69 this is very large and is an impossible task since in this case this value is not the usual basic reproduction number but rather, there is time delay effect and so there should be a correction factor in terms of *I*(*t* − *d_1_*)*/I*(*t* − *d*_2_) multiplying *βS*_0_*/γ*. To find the exact ratio of infected terms, might require stability analysis at equilibrium points or by the next-generation matrix method, see Xu and Ma (2009). Linear or nonlinear analysis is beyond the scope of our present work as here we aim to proposes the model concept. If the present model can reasonably represent the change of infected and recovered individuals, this will motivate deeper analysis in future work.

**Figure 4:**
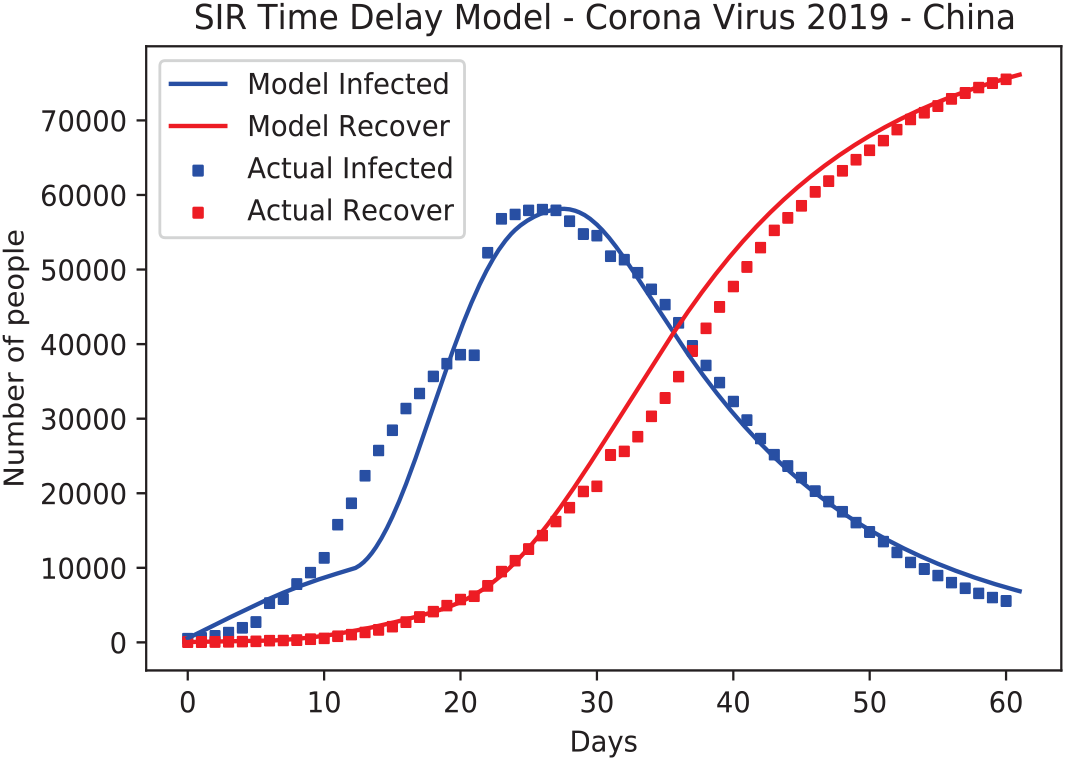
The results of SIR with time Delay model

An interesting representation from the SIR model with the two time delay included is the infected ratio defined by *I*(*t − d*_1_)*/I*(*t − d*_2_); here, we assume that *d*_1_ and *d*_2_ are two integers referring to the number of previous days back from the present day, *t* respectively. From equation (5), we see that *dI/dt >* 0 when

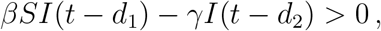

or initially at the early stage of disease spread, 0 *< S* = *S*_0_ < 1,

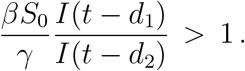

Here, the ratio *R_I_ = I*(*t − d*_1_)*/I*(*t − d*_2_) *>* 0 is a non-dimensional factor multiplying the basic reproduction number *R* = [βS*_0_/γ in the standard SIR model. For the disease spreading period, *I*(*t*) is an increasing function, so *dI/dt >* 0 and *I*(*t − d*_1_) *< I*(*t − d*_2_). When *R_I_* ≪ 1 and 0 *< d*_2_ *< d*_1_, the disease spreads rapidly. Thus, in another way, this shows disease spreading control. The performance of disease control increases when *R_I_* increases. We can use this ratio as an indicator to monitor the progress of disease spread. A plot of the ratio *I* (*t − d*_1_)*/I*(*t − d*_2_) for China during January 22, 2020 to March 15, 2020 is shown in Figure 5 and reveals that China successfully controlled the spread of COVID-19 during this period since *R_I_* (which is actually a function of time) increases monotonically and reaches a steady value at nearly the endemic state; *R_I_* is approaching an asymptotic constant as *t → ∞*. Next, we define a reproduction number 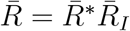 where 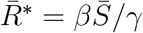, and 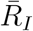 is the average of the ratio *R_I_*. From the simulation over 49 days (61-12 days), we obtain 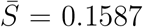 and 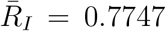, then 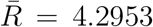. So, we conclude that the basic reproduction number of the COVID-19 spread in China is approximately 4.2953. A more precise closed form with a better estimation of the infected ratio is needed to derive a more accurate reproduction number. It should be noted that all parameters obtained here are from fitting with the data from China. For other countries, all parameters in the model need to be calibrated including the delay times *d*_1_ and d_2_.

**Figure 5:**
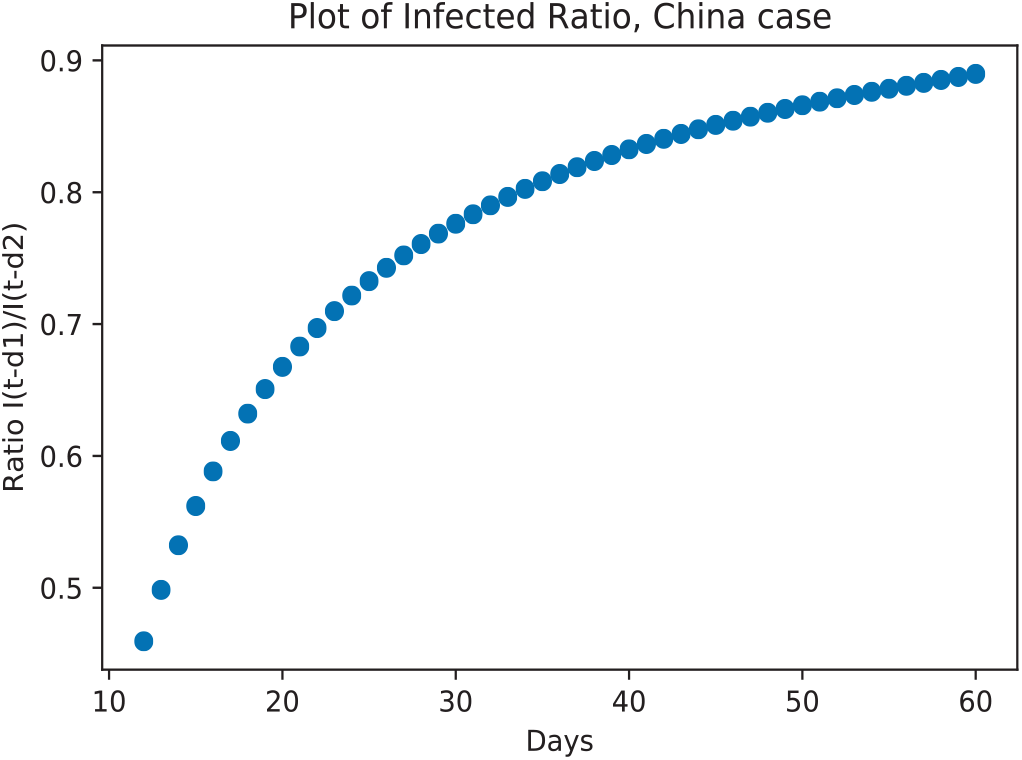
The ratio of infected people *I*(*t − d*_1_)*/I*(*t − d*_2_) over 61 days where *d*_1_ = 12 and *d*_2_ = 5.

## 4 Conclusions

This work presents the SIR model with the addition of a two-time delay for infective individuals. The aim is to extend the standard SIR model to represent the spread of a disease that is spreading rapidly and is readily infectious but does not show symptom during the incubation period, as is the case with the COVID-19. To find the most important rates of contact and recovery, Monte Carlo simulation is performed, running the simulation for the Chinese case since there is information available on all of three stages, starting from early infection growth, rapid infection growth and then to near zero infected cases. Thus, we can determine accurately the number of susceptible individuals in a closed system. Even for the best case of fitting the simulation result to the actual data over 61 days, the standard SIR model gives the basic reproduction number *R\** ≈ 7.61 which is clearly an overestimate. The graph of infected individuals does not fit the actual data well, showing some effects in timing or rate of infected cases. This motivates considering time delay effects in the standard SIR model by inserting two time delays *d*_1_ and *d*_2_ for increasing and decreasing infected individuals respectively.

The results of the SIR model with two-time delays fit well with the actual data and the approximated basic reproduction number is *R\** ≈ 4.2953 where *d*_1_ ≈ 12 and *d*_2_ ≈ 5. The ratio of infected individuals *I*(*t − d*_1_)*/I*(*t − d*_2_) is interesting since it shows the performance of controlling the disease spread using measures such as social distancing or a lock-down policy in each country. The larger the value of the infected ratio, the better the performance of the controls. In the endemic state, the infected ratio approaches an asymptotic constant. We have averaged the infected ratio over the remaining time and applied it as a corrected factor multiplying the usual basic reproduction number to give *R\** ≈ 4.2953 that is more reasonable when compared with influenza *R\** 3 − 4. However, the value of *R\** is different for other countries but we can use this approach to estimate it. Since we have estimated the infected ratio in this work, the closed form formula can be derived using stability analysis at equilibrium points and that is in progress of future reporting.

## Data Availability

https://github.com/CSSEGISandData/COVID-19

## References

Anderson, R. M., and May, R. M. (1979). Population biology of infectious diseases: part i. Nature, 280: 361–367.

Anderson, R. M., and May, R. M. (1998). Infectious diseases of humans: Dynamics and control. Oxford University Press.

Diekmann, O. and Heesterbeek, J. A. P. (2000). Mathematical epidemiology of infectious disease. John Wiley and Sons UK.

Keeling, M. J., and Rohani, P. (2008). Modelling infectious diseases in humans and animals. Princeton University Press.

Kermack, W. and McKendrick, A. (1927). A contribution to the mathematical theory of epidemics. Proceedin of the Royal Society. A, 115: 700–721.

Nadu, C. T. (2020). Novel corona virus 2019 dataset. https://www.kaggle.com/sudalairajkumar/novel-corona-virus-2019-dataset.

Xu, R. and Ma, Z. (2009). Stability of a delayed sirs epidemic model with a nonlinear incidece rate. Chaos, Soliton and Fractals, 41: 2319–2325.

Zulko (2019). Scipy-based delay differential equation (dde) solver. https://pypi.org/project/ddeint.

